# Self-reported stigma and other challenges to HIV treatment adherence for clients in their first 6 months after treatment initiation in Zambia

**DOI:** 10.1101/2025.08.17.25333875

**Authors:** Nancy Scott, Aniset Kamanga, Allison Morgan, Linda Sande, Taurai Makwalu, Kari Radoff, Anushka Reddy Marri, Prudence Haimbe, Priscilla Lumano-Mulenga, Hilda Shakwelele, Sydney Rosen

## Abstract

**Background:** The first six months after initiating or re-initiating antiretroviral therapy (ART) for HIV is the highest risk period for disengagement from care. Little qualitative research has explored ART clients’ experiences in this period in the era of universal ART eligibility. We aimed to understand clients’ challenges in the early treatment period in order to identify potential interventions that could both meet ART clients’ needs and optimize efficiency in this time of declining resources.

**Methods:** We surveyed adults who were initiating or had recently initiated (<6 months) ART at 12 healthcare facilities in two provinces of Zambia. A subset of survey respondents who had experienced or anticipated experiencing adherence challenges were recruited to participate in focus group discussions (FGDs) up to 12 months after the survey. FGDs included up to 10 participants each and were led by facilitators trained in qualitative methods.

**Results:** Fifteen FGDs were conducted (n=126 participants, median age 35). Themes emerged at the individual, interpersonal, facility, and community levels. Stigma related to HIV was the most common theme at all levels. Participants expressed self-stigma, fear of disclosure to family and acquaintances, concerns about what would happen to romantic relationships if their partner learned of their status, and fear of community gossip. Food insecurity was common. Clinic congestion, lack of privacy, and unsympathetic providers emerged as facility-level barriers. Participants mentioned lack of transport money and work conflicts as other impediments. Religious leaders and reliance on traditional healers discouraged ongoing engagement in care.

**Discussion:** Zambian ART clients faced multifaceted barriers to remaining in care during the critical first few months, with particular concern about the insidious roles of stigma influencing their behavior. While societal stigma remains stubbornly difficult to offset, clients’ early clinic experiences—often marked by unwelcome disclosure, facility inefficiency that is perceived as disrespect, and negative provider interactions—and financial and logistical challenges may be amenable to improvement by adapting existing evidence-based interventions.

## Introduction

Achieving continuous and lifelong engagement in HIV treatment for people living with HIV is a critical aspect of global efforts to combat the HIV/AIDS pandemic. The early treatment period, generally defined as the first six months after newly initiating antiretroviral therapy (ART) or re-initiating ART after disengagement(1), is the highest risk interval for both temporary treatment interruptions and long-term disengagement from care. In Zambia, we estimated that 33% of initiating clients had disengaged from care by 6 months, and only 51% remained continuously in care, without any interruptions >28 days, during that period(2). Health system advances that have been demonstrated to improve long-term engagement, such as differentiated service delivery models(3), are generally not available to clients in the early treatment period, as eligibility usually requires at least 6 months of experience on ART and documented viral load suppression(4,5). For the most part, health systems continue to rely on traditional approaches such as requiring frequent clinic visits and adherence counseling sessions to promote engagement in the first six months(6).

The high attrition from care seen during the first six months after starting ART suggests that clients face more challenges or different barriers during this period than later in their treatment journeys. Little qualitative research has explored clients’ experiences in the months immediately after initiation in the era of universal ART eligibility and frequent re-engagement, however(7). A systematic review of the qualitative literature on long-term retention in ART programs in Africa identified poverty and unpredictable life events, self- and social-identity, a “punishing and uninviting” healthcare system, and the challenge of daily medication adherence as the major reasons for disengagement from care during the pre-universal test-and-treat era, when most ART clients had started treatment with more advanced illness and were ART-naïve(8).

In the current universal treatment era, all individuals with HIV are eligible for and encouraged to start ART as quickly as possible, and a majority of those who present for initiation are treatment experienced(9). As both treatment guidelines and the population initiating treatment have changed, it is reasonable to expect client experiences and barriers to engagement to have evolved with them. In an era of funding reductions for global HIV programming, moreover, it is more critical than ever to increase facilities’ operational efficiency by helping them to identify effective interventions to sustain and improve early retention in care (10). By pinpointing effective practices and leveraging proven strategies, HIV programs have an opportunity to optimize resources, minimize service disruptions, and maintain progress toward critical treatment targets[10].

Qualitative inquiry is an effective tool to explore lived experiences and nuanced concepts such as stigma, discrimination, coping mechanisms, and social support networks. We qualitatively explored the barriers to remaining on treatment among people living with HIV in the first six months after initiation in Zambia, using the client perspective to reveal changes to the barriers they face during the early treatment period and to identify opportunities to improve retention and continuity of care to foster program sustainability.

## Methods

### Study design and setting

PREFER-Zambia was a sequential, mixed-methods study conducted as part of the Retain6 project, which aims to identify strategies to improve outcomes during the first six months on ART in Zambia and South Africa. From September 2022 to April 2023, PREFER quantitatively surveyed adults who were initiating, re-initiating, or had recently initiated (<6 months) ART at 12 healthcare facilities in Zambia. PREFER study sites, which were located in Lusaka and Central Provinces and were purposively selected based on ART patient volume, setting (rural and urban), and the differentiated service delivery models offered, are described in previous publications(11,12). ART is provided free of charge at Zambian clinics.

### Sampling and recruitment

Focus group discussions (FGDs) were conducted at 7 of the 12 PREFER study facilities in Zambia, selected to allow sufficient FGD enrollment and represent both urban and rural areas. The FGD study sites included three facilities located in Lusaka Province, consisting of two first level hospitals and one rural health clinic, and four in Central Province, consisting of two district hospitals situated in a peri-urban environment and two urban health centers. We aimed to conduct a total of 15 FGDs with 8-10 participants per group, the number needed to reach saturation, a measure of qualitative validity (13).

Consent for PREFER survey participation included agreement to be contacted up to 12 months after survey enrollment if we wished to invite participants to qualitative FGD. Potential FGD participants were identified from the full PREFER survey sample based on their quantitative survey responses to questions about experiencing or expecting to experience challenges in remaining on HIV treatment. This included those who responded that they had missed any scheduled visits in the past year by 2-3 days or more; expected having difficulty in taking their medications every day or picking up their medication supplies; or reported that the care that they received in the clinic was worse than they expected it to be. Once all potential FGD participants had been identified from the overall PREFER sample, the subsample was stratified by sex to allow for single-sex focus groups to be conducted. The final sample in each facility-based FGD depended on the number of potential participants from that facility. At most sites, females were randomly selected from the list of potential FGD participants, while the full list was used for males, of whom fewer had been enrolled in the PREFER quantitative survey. Participants were then contacted by their preferred mode (telephone or email) and invited to participate.

### Data collection and management

Before commencing each FGD, all participants were briefly screened to re-confirm eligibility and undergo a second informed consent process, specific to the FGD activity. We re-captured participants’ demographic characteristics and assigned the same PREFER unique study ID to allow linkage of demographic data from the quantitative survey.

FGDs were led by two study staff conversant in both English and the appropriate local languages and trained in human subjects’ protection and FDG methodologies. One study team member took notes and one led the FGD using a discussion guide developed from specific issues or concerns that participants brought forward in the quantitative survey (Supplementary file 1). Interview questions focused on facilitators of and barriers to starting and adhering to HIV treatment in the first 6 months of care. All FGDs were single-sex per the guidance of the in-country principal investigator [AK] that separating by sex would be culturally appropriate, create a more comfortable environment, and allow for more open responses. FGDs lasted approximately two hours. Participants received transport reimbursement and a lunch allowance equivalent to US $6.50 (170 ZMW). All FDGs were audio recorded and directly transcribed and translated to English for analysis.

### Data quality and reflexivity statement

To mitigate potential biases brought by the investigators and data collectors, the study team members established trust and rapport with participants before commencing the FGD by transparently identifying their motives and the study goal. Data collectors were trained by study investigators [JK, NS] in qualitative methods, to be active listeners, and to probe in ways that validated participants’ experiences. The local principal investigator [AK] provided oversight and support to data collectors in the field. To ensure data quality, transcripts from the first two FGDs were reviewed for thoroughness and completeness by two study investigators [NS, JK]. The coding and analysis were conducted by a study investigator [NS] and research fellow [KR] and results were interpreted with input from all study authors.

### Analysis and interpretation

For the analysis, we first described aggregate demographic characteristics of FGD participants using linked quantitative survey data. The qualitative codebook was developed using an inductive-deductive approach, wherein most codes were identified jointly by the study team [KR, NS] *a priori* and aligned with the FGD guide and literature but additional codes were allowed to emerge to form a final analytical codebook. Both coders [KR, NS] coded an initial set of transcripts and compared coding. Discrepancies were discussed; one coder [KR] then completed the coding. We then conducted a content analysis in which emerging themes were grouped into larger themes(14). Data were reviewed to ensure that emerging themes were not driven by individual FGDs; only those that emerged across multiple or all groups were considered as key emerging themes.

Emerging themes were organized using the Social-Ecological Model (SEM) framework, which situates individuals within layers of influence, from personal factors to broader societal and environmental factors(15). With the broad understanding that all barriers are interconnected, we adapted the SEM slightly and organized results as barriers experienced by clients at the individual, interpersonal, health facility, and community levels (Fig 1). The SEM framework allowed us to better understand the multi-layered and interconnected barriers that clients experience within the first six months on treatment and provided a structure in which to identify opportunities for intervention. Themes were similar across male and female FGDs, so results were not stratified by sex, but differences are noted when the emergent theme was driven predominantly by one sex. Results are presented in aggregate form, with supporting illustrative quotations when appropriate.

**Fig 1.**
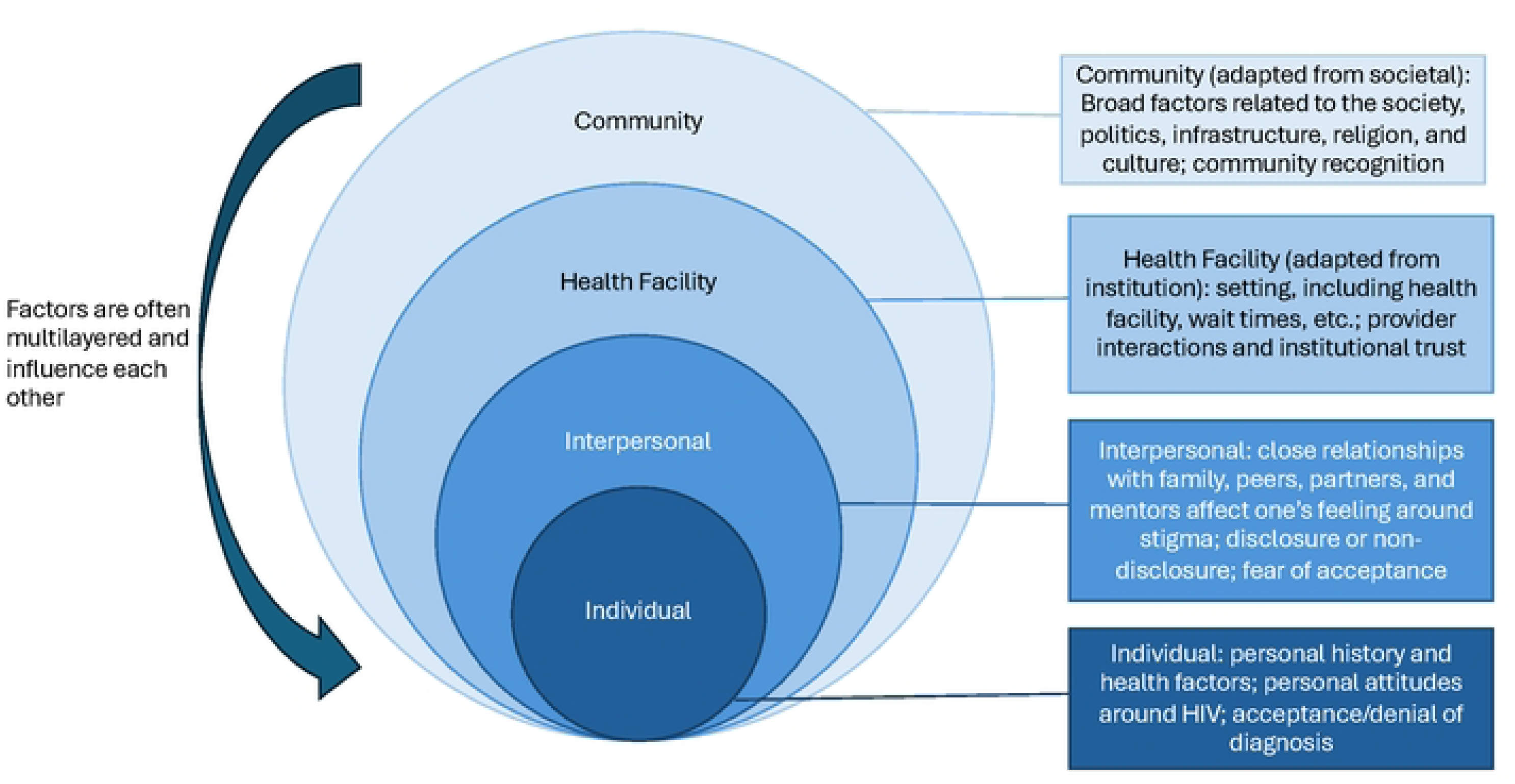
Social Ecological Model Framework as applied to this analysis.

In the results, we briefly describe description of participants’ characteristics, then present emerging barriers by SEM domain. In the discussion, to enhance the interpretation and actionability of our qualitative findings, we mapped key barriers identified through the socioecological model onto the COM-B model (capability, opportunity, and motivation for behavior change) (16). Using the COM-B framework allowed us to identify specific behavioral drivers influencing care-seeking in the first six months of ART, and thus potential areas for targeted, evidence-based intervention.

### Ethical considerations

Ethics approval was obtained from the Boston University Medical Campus IRB (H-42903) in the United States; ERES Converge IRB (2022-June-007) and National Health Research Authority (NHRA000007/10/07/2022) in Zambia; and the Wits University Human Research Ethics Committee (M220535) in South Africa. Written informed consent was obtained via signature or a thumbprint from all participants. FGDs were conducted in private clinical or community settings, and the importance of respect and confidentiality was reviewed with all participants prior to commencing the FGD. Participants were linked to the PREFER survey results using their unique study ID only. Only study staff essential to the data processing had access to the data. PREFER-Zambia was registered at Clinicaltrials.gov as NCT05454852.

## Results

### Characteristics of participants

Fifteen focus groups were conducted with a total of 126 participants. Demographic characteristics reflect the status of the participants at the time of enrollment in the PREFER study, up to 12 months prior to participating in the FGDs (Table 1). Participants had a median age of 35 years; just over half were female. Nearly half (46%) reported that they were newly initiating ART or within one month of initiating when they enrolled in the study; the rest had been on ART up to 6 months at enrollment. About 57% reported food insecurity (participants or other household members sometimes or often going without food) and nearly 88% reported that they would find it difficult or very difficult to access the equivalent of US$4 (100 Zambian Kwacha) if needed for health-related expenses, such as transport to a clinic.

**Table 1:**
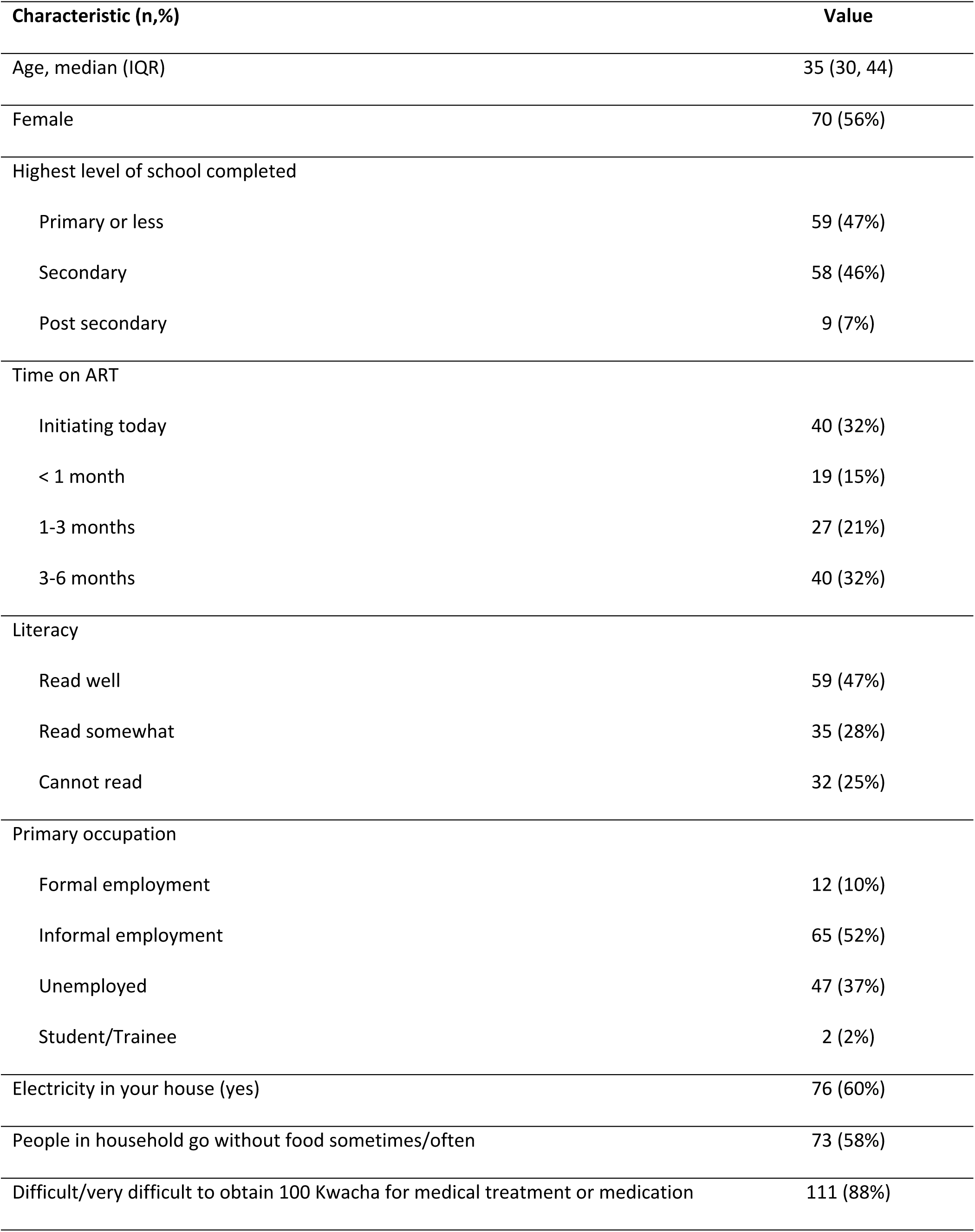
Demographic characteristics of FGD participants in Zambia (n=126)

Qualitative results revealed important insights into challenges faced by clients in the first 6 months of HIV treatment. Structural and social barriers that emerged included food insecurity, stigma at all levels of the SEM, transportation, work conflicts, and a lack of family support. Some facility-level barriers were mentioned but were not a major area of a concern for participants.

### Individual and interpersonal levels

Individual and interpersonal barriers to remaining in care are detailed in Table 2. At the individual level, the theme that emerged most strongly was self-stigma, or the internalization of negative stereotypes and social prejudices. Participants discussed the challenges of confronting and accepting their diagnosis and adhering to daily medication therapy. Participants here also reported vacillating between acceptance and denial, potentially leading to inconsistent treatment engagement. Food insecurity also emerged as a barrier at the individual level, reflecting the high proportion of participants (57%) who reported going without food sometimes or often. Participants reported increases in appetite when initiating treatment and had insufficient food. Side effects and missing doses were also attributed to not having food.

**Table 2.** Client reported individual- and interpersonal-level challenges to HIV adherence in the early treatment period.

| Level of SEM | Theme | Sub-theme | Illustrative quotes |
| --- | --- | --- | --- |
| Individual | Stigma | Internalized stigma and difficulty | “When starting medication, the biggest challenge people pass through is acceptance that today I start medication, it is difficult. Many people that is a challenge, even us who are here; one side you accept, the other side |
|  |  | accepting diagnosis | it's denial. You feel like you are dreaming, looking for the truth. Until at last you accept."–*female* |
|  |  |  | "Just from the beginning you get filled with thoughts like you should just stop because people are laughing at you, so it is better to stop and die." - *female* |
|  | Food insecurity | Increased appetite from medication<br>Taking medication on empty stomach | "I found a challenge with eating. Wanting to eat very much but not having something to eat."–*male*<br>"At the beginning when you start medication when you don't have money it's a problem because the drug demands you to eat hence you forget to take medication because of lack of food saying I will drink [the medication] after I eat and you forget and [you miss a dose]."– *female*<br>"We said you can't take the drugs without eating which makes you feel sick. But you wake up again and take them because that's your life." – *female* |
| Interpersonal | Stigma and non-disclosure | Female clients fear loss of romantic relationships or marriages | "Today I am a single lady if I find a man he would want to marry me, he will ask about my status. I will answer him that I don't take medication. Because I like love, I will surely stop taking medicine. I will hide."– *female*<br>"A lot of people hide their HIV status just because they want to be in a relationship. So, eventually instead of taking treatment they stop, and we have seen some people have died because of that, because you don't want your partner to know that you are taking HIV drugs"– *female* |
|  |  |  | “Because us ladies we always want to hide ourselves. I don’t want my boyfriend to know that am sick so I just stop so that I continue with my boyfriend. That becomes the greatest challenge.” *–female*<br>“Some find that you get married the husband is okay, he doesn’t take medication, so they hide. They choose marriage over their health. They say let me stop before I am discovered.” *–female*<br>“Now you are married how are you going to tell your partner that you are taking medicine because most of us ladies we tend to say because he doesn’t know let me just stop. In fear to lose someone you end up denying taking your medicine because you want to please that person so at the end of the day, most of them they stop... In fear of [losing him] you end up dropping the medicine, ending in problems.” *–female*<br>“That person can take HIV drugs for some time even for one year or two years and then when that person finds some to marry, that person will decide to stop taking the HIV drugs. What that person needs to do is to open up but it’s not easy and people tend to hide their HIV status from their partners. That’s the biggest reason people start and stop HIV treatment” *–male*<br>“My neighbor is on treatment and a man wanted to marry her; she never disclosed her status to him. When the man got her, she stopped taking the drugs. She stayed for 6 months without taking them. At that point she got very sick.” *–female* |
|  | Social support | • Presence or absence of family/friend support influences early adherence | “My relative also used to speak ill of me that I was left with the virus, people talk as if it was deliberate to be infected...If you are not strong and you do not have a counsellor to help you, you can kill yourself.” – *female*<br>“This issue of the family laughing at us is the challenge. They started blaming my husband for infecting me with the virus and he never cared. I was told to leave him, of which I did not...they would gossip about me but they never had the guts to tell me in my face.” – *female*<br>“I think for me what main reason that causes people to start and stop the HIV treatment is they fear being laughed at by their friends. With some people you find that they even throw away their HIV drugs along the road because they fear being laughed at.” – *male* |

At the interpersonal level, stigma was also the dominant theme, particularly as it relates to disclosure within intimate relationships. Participants from nearly all of our female-only FGDs described the reactions and perceptions of those with whom they are in close relationships or the prospect of romantic relationships as factors affecting ART continuation. Non-disclosure of HIV status was frequently reported and is often associated with stigma and a fear of abandonment. Participants described that fear of unwanted disclosure in close relationships, including marriages, leads to hiding behaviors, reticence about care seeking, and sometimes, treatment interruption.

In addition, social support, both positive and negative, emerged as an important theme for clients during the first 6 months of ART. Perceived lack of support and intra-familial stigma were reported as negatively impacting adherence, while, conversely, positive support from family and friends was described as motivating for ART adherence. Participants reported, for example:

> *“My family even knows what time I take the medication, so they are always encouraging me to take the HIV drugs at the right time.” -male*
>
> *“If you have friends and explain the problem to them, friends can encourage and help give strength, rather than being alone which can cause heartbreak.” –female Facility level*

At the facility level, three themes arose: anticipated stigma, lack of respect by healthcare providers, and inefficient clinic processes leading to long waiting times (Table 3). Participants feared non-consensual disclosure of their status and subsequent experience of stigma. They explained that many people feel anxious that they may encounter people that they know when attending clinic, who in turn may disclose their status without their consent.

**Table 3.**
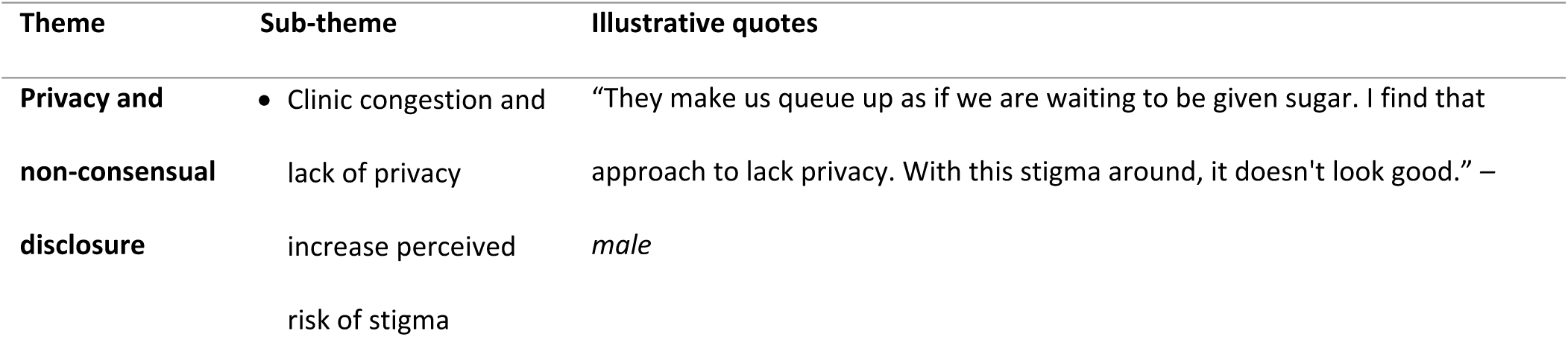
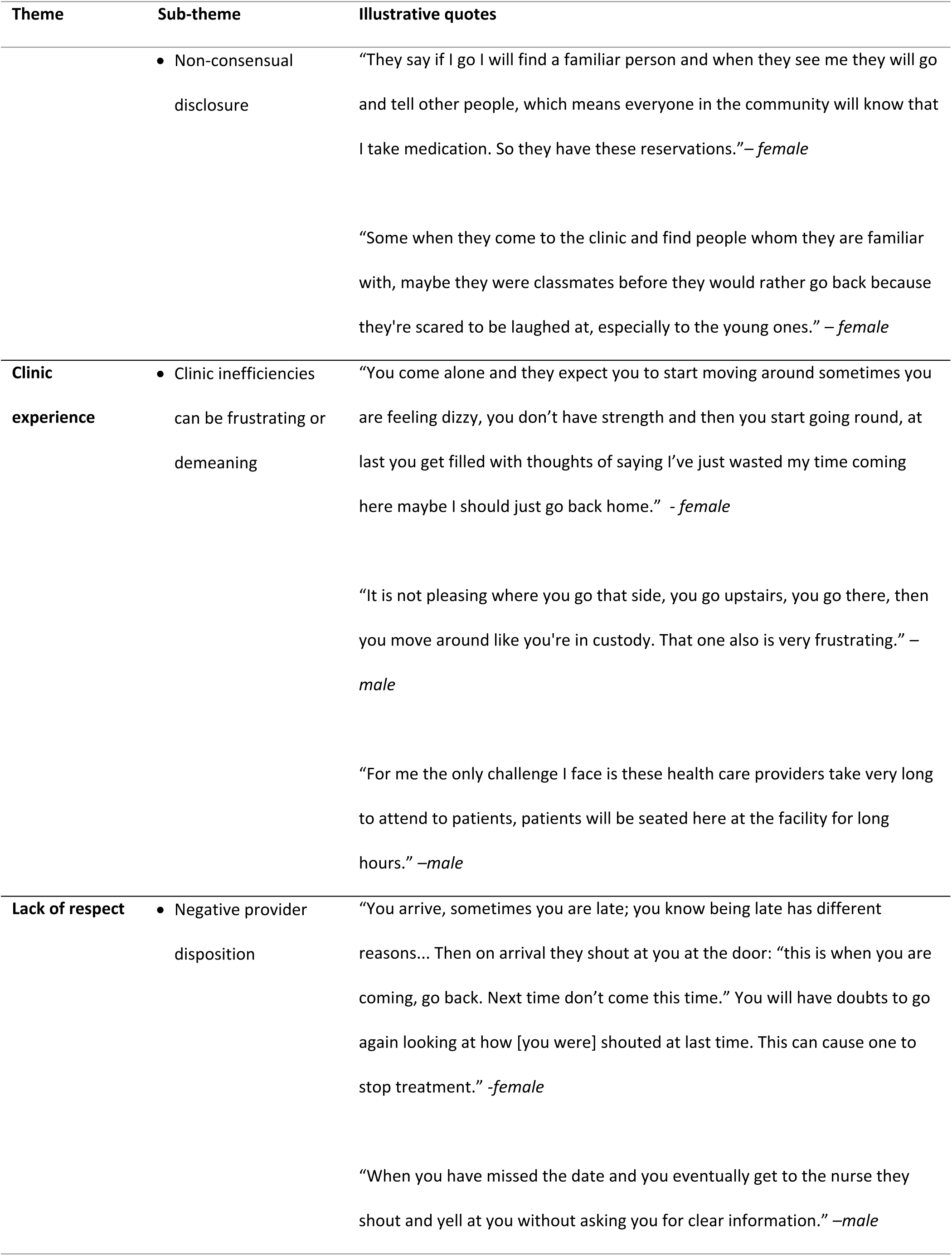
Client reported facility-level challenges to HIV adherence in the early treatment period.

In describing providers’ lack of respect, participants used language that suggested a perception of being treated like children or prisoners. Participants also explained that having a negative encounter with a provider can discourage remaining in care. They desired compassion and understanding, especially with missed appointments, and wanted to be attended to by providers with kind, inquisitive dispositions.

### Community level

Stigma also emerged as a salient theme at the community level, alongside scheduling conflicts, transportation challenges, and religion and traditional culture (Table 4). Clients described experiencing and anticipating ridicule, criticism, and gossip and expressed fear of being singled out by community members. They explained that some people choose to bypass nearby clinics to avoid disclosure and gossip within the community, which can contribute to increased transportation and opportunity costs as they travel to more distant facilities.

**Table 4.**
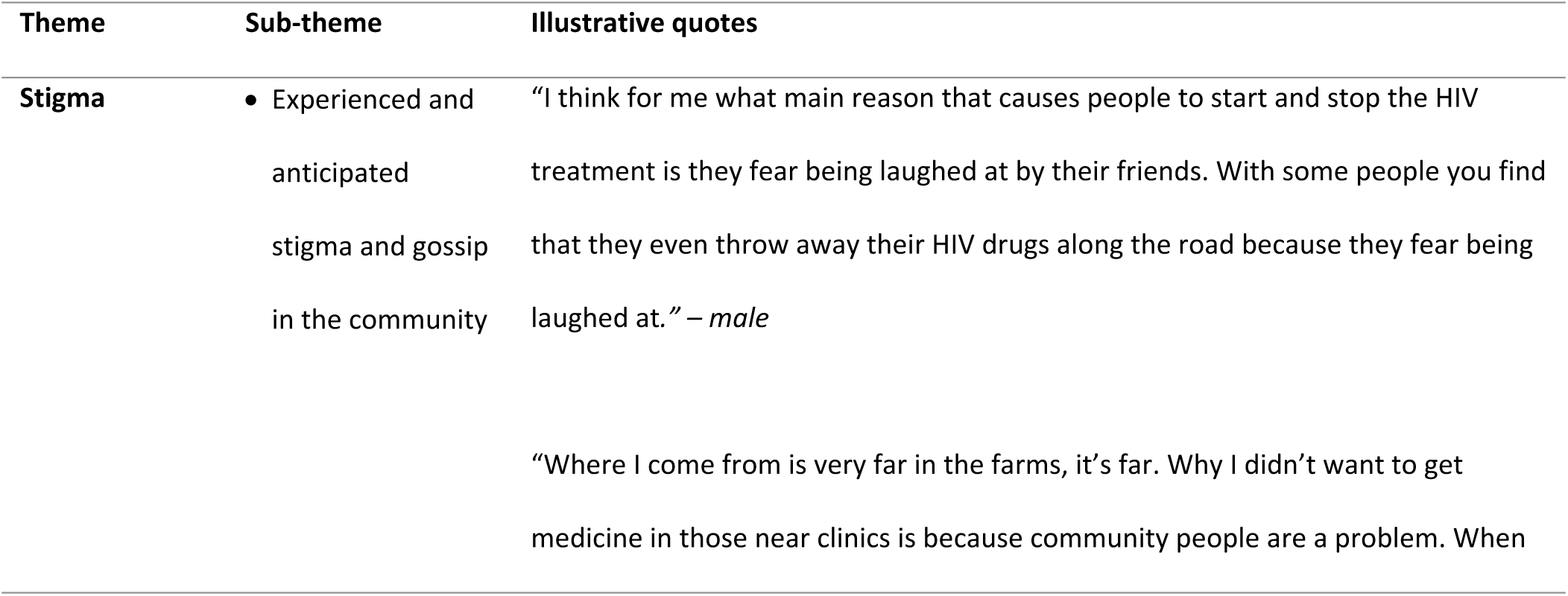
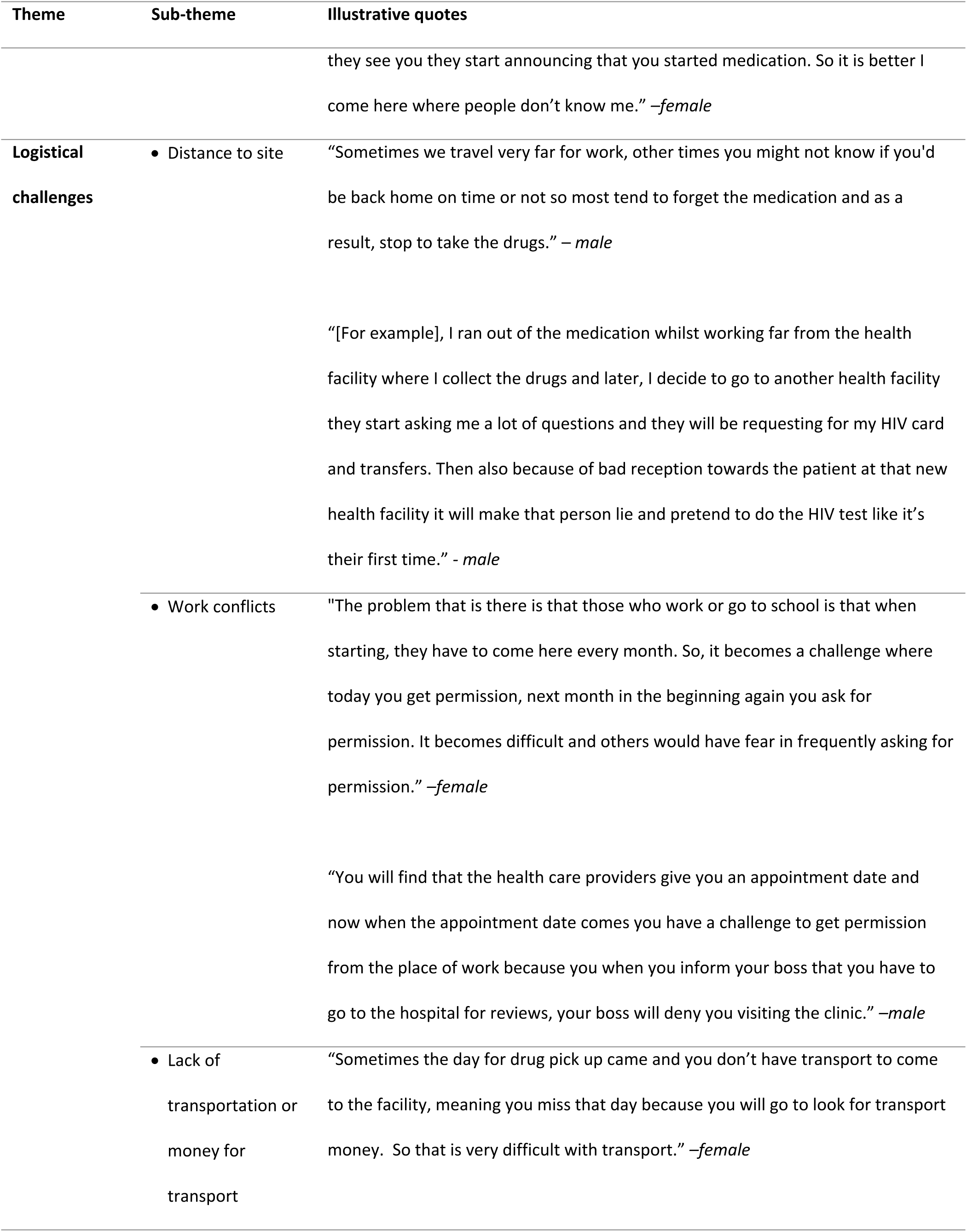
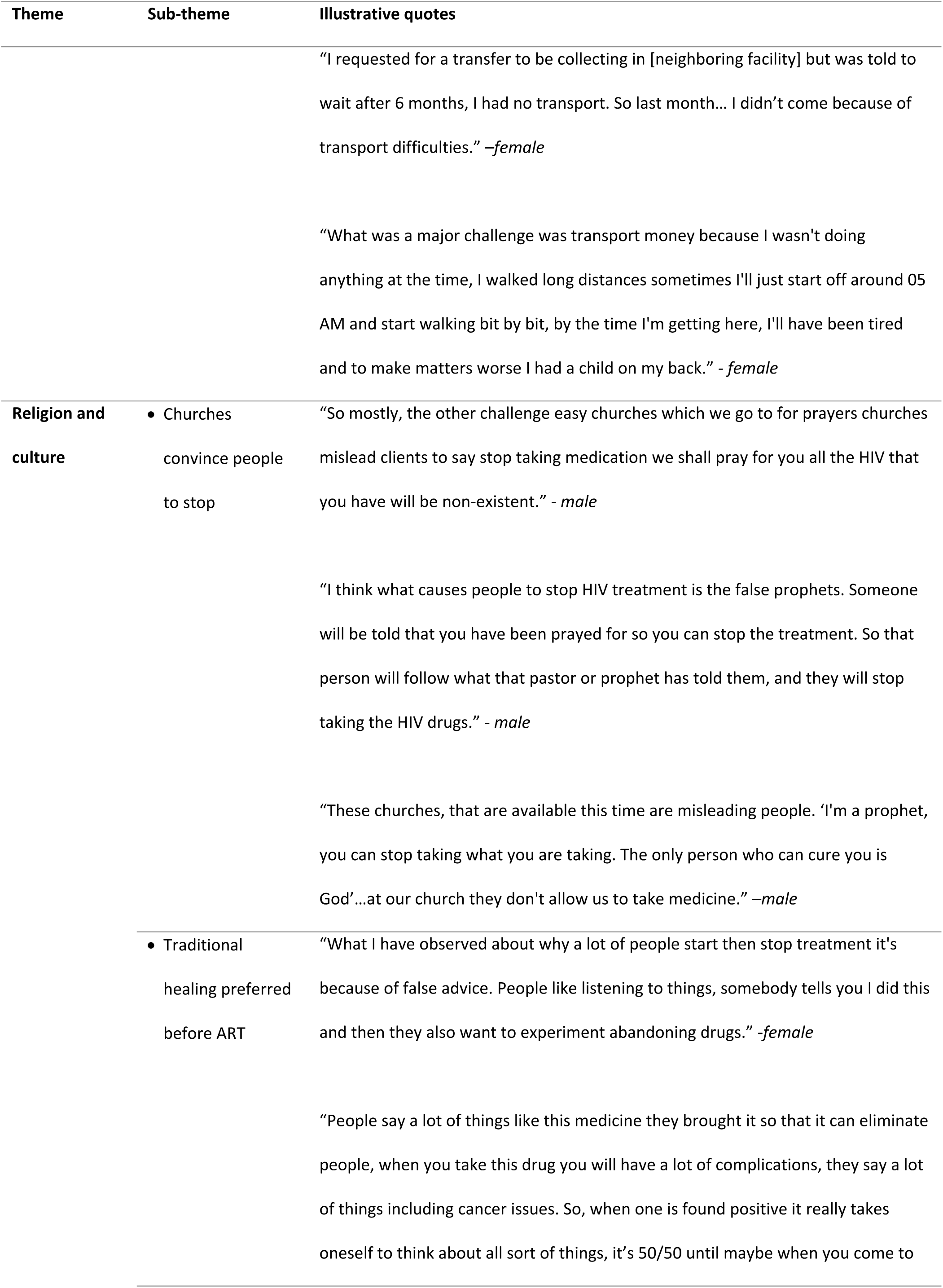
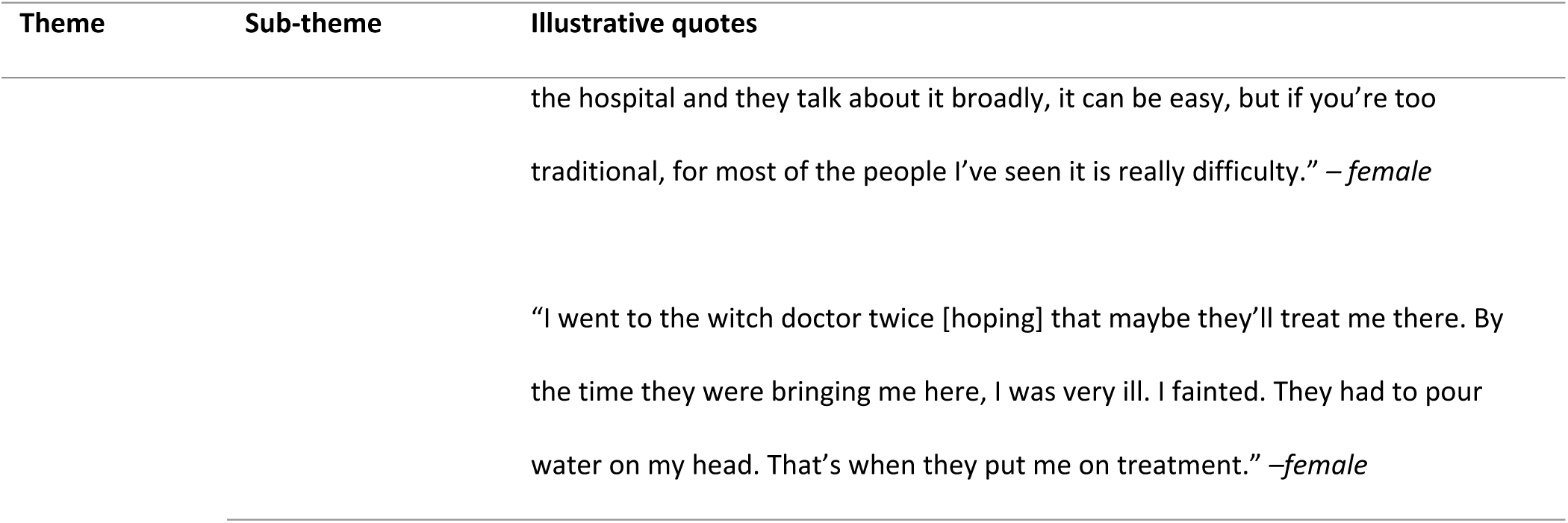
Client reported community-level challenges to HIV adherence in the early treatment period.

Many clients reported encountering logistical difficulties with work schedules in the initial 6 months following treatment initiation, which is a period when frequent clinic visits are required. Many feared disclosure of their status and job loss if they request time off to attend clinic visits. Some struggled to pick up medications due to their work or school commitments and being denied time off. Others found it challenging to adhere to scheduled clinic visits, especially if they travel for work. Clients reported that for many of those who travel for work, though not all, it can be easier to stop completely or to present at a different facility as a ‘new’ patients than to navigate the transfer process. When discussing costs of seeking care, lack of funding for transportation was raised as a concern, though participants generally agreed that costs incurred at the facility itself are not a barrier to early adherence, as ART is given free of charge.

Lastly, religion and culture emerged as an important theme that influenced treatment retention during the early treatment period. Participants in many of the FGDs explained that many prophets, pastors, and online churches promote prayer to cure HIV instead of, not in conjunction with, ART. Participants explained that many people are also encouraged to first or exclusively utilize traditional curative practices, mentioning visiting witch doctors, taking herbs, or drinking the fat of a crocodile as traditional remedies for HIV.

## Discussion

Our study contributes uniquely to the existing literature by specifically examining the barriers and facilitators experienced during the critical six-month period following ART initiation, an interval which remains understudied despite its recognized importance for long-term treatment success (6). We qualitatively explored the challenges and lived experiences of clients in the first six months after initiating or re-initiating ART in two provinces in Zambia. We found that, despite the more than two decades since large-scale treatment programs were launched and the near universality of treatment access in countries like Zambia, stigma remains a potent obstacle at all levels of the SEM. Participants described stigma as both internalized stigma (experiencing self-judgment) and as external stigma (anticipating or experiencing judgment from others within their close relationships, at facility visits, and at various interactions within communities). At the individual level, the fear of being judged or rejected if one’s HIV status is disclosed led to intentional non-disclosure, often disrupting treatment. This is particularly evident in the interpersonal domain, where non-disclosure was driven by the desire to preserve romantic relationships or avoid conflict within marriages. These findings, primarily reported by women, align with other research in similar contexts where women report internalized stigma and non-disclosure of HIV status as barriers that influence care seeking and treatment adherence (17,18)[19]. At the community-level, both our findings and other studies in sub-Saharan Africa highlight the pervasive influence of community stigma on ART retention (19,20).

A 2017 review identified stigma as the third most frequently patient-reported barrier to ART adherence in sub-Saharan Africa, preceded only by forgetfulness and food insecurity(21). While it is well documented that stigma can negatively affect HIV outcomes and continued engagement in care(22–24), we found only one longitudinal study in the literature --conducted among patients in the United States-- that examined the effects of experienced and perceived stigma during the early treatment period on treatment outcomes(25). It concluded that internalized stigma and depression in the early treatment period mediate HIV outcomes, including viral load suppression. While stigma remains stubbornly persistent and food insecurity a pervasive challenge, it is noteworthy that previously dominant barriers such as concerns about side effects, misconceptions regarding ART, and misinformation about treatment duration (26) were rarely mentioned by our participants. This suggests substantial progress in patient education and drug tolerability, reflecting important advancements in ART delivery.

In an era marked by diminishing funding for global HIV programming, identifying efficient interventions and strategically adapting them to address early retention challenges is critical. Responding directly to barriers identified by at-risk patients, such as stigma, food insecurity, and lack of psychosocial support, may help facilities target interventions to those at greatest need and thus improve resource allocation. To make our findings more practical and actionable for providers, we applied the COM-B model (capability, opportunities, motivation for behavior change) to our results to pinpoint the behavioral drivers and identify evidence-based interventions that may be adapted for use among clients in the early treatment period.

**Capability**, which includes both psychological (knowledge, resilience) and physical (health, physical skills) components, aligns closely with barriers at the individual level. Psychological capability was particularly affected by internalized stigma, resulting in ambivalence about diagnosis acceptance. While acceptance is a pivotal factor in motivating participants to initiate and sustain treatment (27), participants described vacillating between acceptance and denial, which could lead to inconsistent adherence. Enhancing psychological capability could involve interventions such as counseling or structured peer-support to bolster mental resilience and self-acceptance post-diagnosis (28). Peer support through expert clients and support groups may also create trusted social networks in which clients can more comfortably discuss fears, disclosure concerns, and overcoming stigma (29,30). Physical capability was challenged by side effects, hunger, and physical symptoms exacerbated by inadequate nutrition. Programs ensuring food supplementation or nutritional support at ART initiation could directly address these physical capability constraints (31,32).

**Opportunity** encompasses both social (interpersonal and community environments) and physical (environmental, logistical, and structural) aspects. Barriers relating to interpersonal and community-level stigma significantly limited social opportunity by restricting social support, increasing fear of disclosure, and complicating interpersonal relationships, especially among women. Interventions here could include community stigma-reduction campaigns, couples’ counseling services, or targeted strategies encouraging safe disclosure within intimate relationships (33,34). Physical opportunity was constrained by facility-level inefficiencies (long wait times, congestion, lack of privacy) and by logistical challenges (transportation costs, scheduling conflicts). Addressing these would require streamlining clinic processes, decentralizing ART services, implementing flexible dispensing schedules, and subsidizing or facilitating transport arrangements during the early treatment period. Many of these options are already offered to stable clients through differentiated service delivery models (35–37) but generally denied to early treatment clients, as mentioned above.

**Motivation**, reflecting both reflective (beliefs about HIV and ART treatment) and automatic (emotional responses to ART and HIV status) components, was influenced by religious and cultural beliefs promoting alternative healing practices, as well as by emotional reactions (fear, anxiety, shame) stemming from stigma and anticipated negative experiences in healthcare settings. Interventions targeting motivation could include engaging religious and traditional leaders in ART education, implementing supportive counseling sessions during initial visits, and promoting positive provider-patient interactions that build trust and comfort, thereby improving early emotional associations with ART (38,39).

Although qualitative research inherently limits generalizability, our rigorous approach including reaching thematic saturation, ensures robust and credible findings unique to those in the early HIV treatment period in Zambia. Our study included only two of Zambia’s ten provinces, however, and may not represent the full range of experiences across the country. Additionally, we recruited participants who were actively seeking treatment and thus did not capture the experiences of individuals who have not engaged with or have disengaged from care. Finally, we recognize that in the context of a FGD, participants may have been reluctant to admit to behaviors or experiences, such as having missed clinic visits in the past, that could incur negative reactions from others in the FGD or the study staff. Further studies should expand geographically and methodologically, particularly engaging clients who have disengaged from care to further inform targeted early-treatment interventions.

## Conclusions

For HIV treatment, as for many other experiences, initial perceptions shape long-term expectations and behaviors (40). We found that ART clients who are new to treatment face multifaceted barriers to remaining in care during the critical first few months, with particular concern about the insidious roles of stigma in influencing their behavior. While societal stigma remains stubbornly difficult to offset, clients’ first-hand clinic experiences—often marked by unwelcome disclosure, inefficiencies, and negative provider interactions—and logistical challenges, such as lack of transport, may be amenable to improvement and prove critical in shaping early engagement in care. Building on patients’ own accounts of the challenges they face has the potential to identify and adapt interventions and strategies that could increase retention in care during the early treatment period and the years to come.

## Data Availability

Data generated by the study will be made publicly available in the Open BU repository (https://open.bu.edu/) after the PREFER study protocol has been closed (anticipated closure December 2026). Qualitative data will be presented in aggregate and coded to avoid any possible identification. Until then, data will remain under the supervision of the Boston University Medical Campus IRB, the University of the Witwatersrand Human Research Ethics Committee (HREC), and the ERES-Converge IRB. Requests can be sent to the BUMC IRB at.

## Acknowledgements

We would like to thank the clients and staff of the study sites for their cooperation in allowing us to conduct this study and the South Africa National Department of Health and Zambian Ministry of Health for approving this research.

## Supplementary files

S1. Focus group discussion guide

## References

1. Ehrenkranz P, Rosen S, Boulle A, Eaton JW, Ford N, Fox MP, et al. The revolving door of HIV care: Revising the service delivery cascade to achieve the UNAIDS 95-95-95 goals. PLoS Med. 2021 May 1;18(5).

2. Benade M, Maskew M, Chilembo P, Mwanza MW, Savory T, Nichols BE, et al. Patterns of engagement in care during clients’ first 12 months after HIV treatment initiation in Zambia: a retrospective cohort analysis using routinely collected data. medRxiv [Internet]. 2024 Oct 4; Available from: http://medrxiv.org/lookup/doi/10.1101/2024.10.03.24314849

3. Long L, Kuchukhidze S, Pascoe S, Nichols BE, Fox MP, Cele R, et al. Retention in care and viral suppression in differentiated service delivery models for HIV treatment delivery in sub-Saharan Africa: a rapid systematic review. J Int AIDS Soc. 2020;23(11):1–14.

4. Prust ML, Banda CK, Nyirenda R, Chimbwandira F, Kalua T, Jahn A, et al. Multi-month prescriptions, fast-track refills, and community ART groups: results from a process evaluation in Malawi on using differentiated models of care to achieve national HIV treatment goals. J Int AIDS Soc. 2017 Jul 21;20(S4).

5. Grimsrud A, Bygrave H, Doherty M, Ehrenkranz P, Ellman T, Ferris R, et al. Reimagining HIV service delivery: the role of differentiated care from prevention to suppression. J Int AIDS Soc [Internet]. 2016 Jan [cited 2024 Jul 24];19(1). Available from: https://onlinelibrary.wiley.com/doi/10.7448/IAS.19.1.21484

6. Rosen S, Grimsrud A, Ehrenkranz P, Katz I. Models of service delivery for optimizing a patient’s first six months on antiretroviral therapy for HIV: an applied research agenda. Gates Open Res [Internet]. 2020 [cited 2024 Aug 12];4:1–15. Available from: /pmc/articles/PMC7445417/

7. Kim MH, Zhou A, Mazenga A, Ahmed S, Markham C, Zomba G, et al. Why Did I Stop? Barriers and Facilitators to Uptake and Adherence to ART in Option B+ HIV Care in Lilongwe, Malawi. PLoS One. 2016 Feb 22;11(2):e0149527.

8. Ware NC, Wyatt M a, Geng EH, Kaaya SF, Agbaji OO, Muyindike WR, et al. Toward an understanding of disengagement from HIV treatment and care in sub-Saharan Africa: A qualitative study. PLoS Med [Internet]. 2013 Jan [cited 2013 Jan 28];10(1):e1001369. Available from: http://www.pubmedcentral.nih.gov/articlerender.fcgi?artid=3541407&tool=pmcentrez&rendert ype=abstract

9. Benade M, Maskew M, Juntunen A, Flynn DB, Rosen S. Prior exposure to antiretroviral therapy among adult patients presenting for HIV treatment initiation or reinitiation in sub-Saharan Africa: a systematic review. BMJ Open [Internet]. 2023 Nov 19;13(11):e071283. Available from: https://bmjopen.bmj.com/lookup/doi/10.1136/bmjopen-2022-071283

10. Grimsrud A, Holmes CB, Sande L. Build, do not dismantle: leveraging a differentiated service delivery approach for broader health impact amidst funding changes. J Int AIDS Soc [Internet]. 2025 Jul 1 [cited 2025 Jul 28];28(S3):e26514. Available from: /doi/pdf/10.1002/jia2.26514

11. Maskew M, Ntjikelane V, Juntunen A, Scott N, Benade M, Sande L, et al. Preferences for services in a patient’s first six months on antiretroviral therapy for HIV in South Africa and Zambia (PREFER): research protocol for a prospective observational cohort study. Gates Open Res. 2024;7.

12. Pascoe S, Huber A, Mokhele I, Lekodeba N, Ntjikelane V, Sande L, et al. The SENTINEL study of differentiated service delivery models for HIV treatment in Malawi, South Africa, and Zambia: research protocol for a prospective cohort study. BMC Health Serv Res. 2023;23(1).

13. Hennink MM, Kaiser BN, Weber MB. What Influences Saturation? Estimating Sample Sizes in Focus Group Research. Qual Health Res. 2019;29(10).

14. Elo S, Kyngäs H. The qualitative content analysis process. J Adv Nurs [Internet]. 2008 Apr 1 [cited 2024 Dec 2];62(1):107–15. Available from: https://onlinelibrary-wiley-com.ezproxy.bu.edu/doi/full/10.1111/j.1365-2648.2007.04569.x

15. Mcleroy KR, Bibeau D, Steckler A, Glanz K. An ecological perspective on health promotion programs. Health Educ Q [Internet]. 1988 [cited 2024 Apr 24];15(4):351–77. Available from: https://pubmed.ncbi.nlm.nih.gov/3068205/

16. Michie S, van Stralen MM, West R. The COM-B Model of Behaviour [Internet]. [cited 2024 Oct 30]. Available from: https://social-change.co.uk/files/02.09.19_COM-B_and_changing_behaviour_.pdf

17. Ziblim AM, Inusah AHS, Boah M. Perceptions of patients and healthcare providers regarding barriers and enablers of HIV anti-retroviral therapy among women at a regional hospital in Ghana: implications for national HIV/AIDS control. BMC Womens Health. 2024 Dec 1;24(1).

18. Alhassan Y, Twimukye A, Malaba T, Myer L, Waitt C, Lamorde M, et al. ‘I fear my partner will abandon me’: the intersection of late initiation of antenatal care in pregnancy and poor ART adherence among women living with HIV in South Africa and Uganda. BMC Pregnancy Childbirth. 2022 Dec 1;22(1).

19. Turan B, Budhwani H, Fazeli PL, Browning WR, Raper JL, Mugavero MJ, et al. How Does Stigma Affect People Living with HIV? The Mediating Roles of Internalized and Anticipated HIV Stigma in the Effects of Perceived Community Stigma on Health and Psychosocial Outcomes. AIDS Behav. 2017 Jan 1;21(1):283–91.

20. Katz IT, Ryu AE, Onuegbu AG, Psaros C, Weiser SD, Bangsberg DR, et al. Impact of HIV-related stigma on treatment adherence: systematic review and meta-synthesis. J Int AIDS Soc. 2013;16(3 Suppl 2).

21. Croome N, Ahluwalia M, Hughes LD, Abas M. Patient-reported barriers and facilitators to antiretroviral adherence in sub-Saharan Africa. Vol. 31, AIDS. Lippincott Williams and Wilkins; 2017. p. 995–1007.

22. Hoffman S, Tymejczyk O, Kulkarni S, Lahuerta M, Gadisa T, Remien RH, et al. Stigma and HIV Care Continuum Outcomes Among Ethiopian Adults Initiating ART [Internet]. 2017. Available from: www.jaids.com

23. Treves-Kagan S, Steward WT, Ntswane L, Haller R, Gilvydis JM, Gulati H, et al. Why increasing availability of ART is not enough: A rapid, community-based study on how HIV-related stigma impacts engagement to care in rural South Africa. BMC Public Health. 2016 Jan 28;16(1).

24. Sweeney SM, Vanable PA. The Association of HIV-Related Stigma to HIV Medication Adherence: A Systematic Review and Synthesis of the Literature. AIDS Behav [Internet]. 2016;20(1):29–50. Available from: 10.1007/s10461-015-1164-1

25. Yigit I, Turan B, Kurt ah, Weiser SD, Johnson MO, Mugavero MJ, et al. Longitudinal Associations of Experienced and Perceived Community Stigma With Antiretroviral Therapy Adherence and Viral Suppression in New-to-Care People With HIV: Mediating Roles of Internalized Stigma and Depression Symptoms [Internet]. 2024. Available from: www.jaids.com

26. Murray LK, Semrau K, McCurley E, Thea DM, Scott N, Mwiya M, et al. Barriers to acceptance and adherence of antiretroviral therapy in urban Zambian women: a qualitative study. AIDS Care [Internet]. 2009 Jan [cited 2014 Mar 25];21(1):78–86. Available from: http://www.pubmedcentral.nih.gov/articlerender.fcgi?artid=2821879&tool=pmcentrez&rendertype=abstract

27. Nam SL, Fielding K, Avalos A, Dickinson D, Gaolathe T, Geissler PW. The relationship of acceptance or denial of HIV-status to antiretroviral adherence among adult HIV patients in urban Botswana. Soc Sci Med [Internet]. 2008 [cited 2025 Aug 10];67(2):301–10. Available from: https://pubmed.ncbi.nlm.nih.gov/18455285/

28. Denison JA, Burke VM, Miti S, Nonyane BAS, Frimpong C, Merrill KG, et al. Project YES! Youth Engaging for Success: A randomized controlled trial assessing the impact of a clinic-based peer mentoring program on viral suppression, adherence and internalized stigma among HIV-positive youth (15-24 years) in Ndola, Zambia. PLoS One [Internet]. 2020 [cited 2025 Jul 28];15(4). Available from: https://pubmed.ncbi.nlm.nih.gov/32240186/

29. Hlongwa M, Cornell M, Malone S, Pitsillides P, Little K, Hasen N, et al. Uptake and Short-Term Retention in HIV Treatment Among Men in South Africa: The Coach Mpilo Pilot Project [Internet]. Available from: www.ghspjournal.org

30. MacKellar D, Williams D, Bhembe B, Dlamini M, Byrd J, Dube L, et al. Peer-Delivered Linkage Case Management and Same-Day ART Initiation for Men and Young Persons with HIV Infection — Eswatini, 2015–2017. Morbidity and Mortality Weekly Report [Internet]. 2018 Jun 15 [cited 2025 Jul 30];67(23):663. Available from: https://pmc.ncbi.nlm.nih.gov/articles/PMC6002033/

31. Cantrell RA, Sinkala M, Megazinni K, Lawson-Marriott S, Washington S, Chi BH, et al. A pilot study of food supplementation to improve adherence to antiretroviral therapy among food insecure adults in Lusaka, Zambia. J Acquir Immune Defic Syndr [Internet]. 2008 Oct [cited 2025 Jul 28];49(2):10.1097/QAI.0b013e31818455d2. Available from: https://pmc.ncbi.nlm.nih.gov/articles/PMC3847664/

32. Nagata JM, Cohen CR, Young SL, Wamuyu C, Armes MN, Otieno BO, et al. Descriptive Characteristics and Health Outcomes of the Food by Prescription Nutrition Supplementation Program for Adults Living with HIV in Nyanza Province, Kenya. PLoS One [Internet]. 2014 Mar 19 [cited 2025 Jul 28];9(3):e91403. Available from: https://journals.plos.org/plosone/article?id=10.1371/journal.pone.0091403

33. Kennedy CE, Haberlen S, Amin A, Baggaley R, Narasimhan M. Safer disclosure of HIV serostatus for women living with HIV who experience or fear violence: A systematic review. J Int AIDS Soc [Internet]. 2015 Dec 1 [cited 2025 Jul 28];18(Suppl 5). Available from: https://pubmed.ncbi.nlm.nih.gov/26643462/

34. Stangl AL, Lloyd JK, Brady LM, Holland CE, Baral S. A systematic review of interventions to reduce HIV-related stigma and discrimination from 2002 to 2013: how far have we come? J Int AIDS Soc [Internet]. 2013 [cited 2025 Jul 28];16(3Suppl 2):18734. Available from: https://pmc.ncbi.nlm.nih.gov/articles/PMC3833106/

35. Grimsrud A, Barnabas R V., Ehrenkranz P, Ford N. Evidence for scale up: the differentiated care research agenda. J Int AIDS Soc [Internet]. 2017 Jul 21 [cited 2025 Jul 28];20(Suppl 4):22024. Available from: https://pmc.ncbi.nlm.nih.gov/articles/PMC5577722/

36. Roy M, Bolton Moore C, Sikazwe I, Holmes CB. A Review of Differentiated Service Delivery for HIV Treatment: Effectiveness, Mechanisms, Targeting, and Scale. Curr HIV/AIDS Rep [Internet]. 2019 Aug 15 [cited 2025 Jul 28];16(4):324–34. Available from: https://pubmed.ncbi.nlm.nih.gov/31230342/

37. Mody A, Roy M, Sikombe K, Savory T, Holmes C, Bolton-Moore C, et al. Improved Retention with 6-Month Clinic Return Intervals for Stable Human Immunodeficiency Virus-Infected Patients in Zambia. Clinical Infectious Diseases [Internet]. 2018 Jan 15 [cited 2025 Jul 28];66(2):237–43. Available from: https://pubmed.ncbi.nlm.nih.gov/29020295/

38. Zou J, Yamanaka Y, John M, Watt M, Ostermann J, Thielman N. Religion and HIV in Tanzania: Influence of religious beliefs on HIV stigma, disclosure, and treatment attitudes. BMC Public Health [Internet]. 2009 Mar 4 [cited 2025 Jul 28];9(1):1–12. Available from: https://bmcpublichealth.biomedcentral.com/articles/10.1186/1471-2458-9-75

39. Campbell C, Scott K, Skovdal M, Madanhire C, Nyamukapa C, Gregson S. A good patient? How notions of “a good patient” affect patient-nurse relationships and ART adherence in Zimbabwe. BMC Infect Dis [Internet]. 2015 Sep 30 [cited 2025 Jul 28];15(1):1–11. Available from: https://bmcinfectdis.biomedcentral.com/articles/10.1186/s12879-015-1139-x

40. Oben P. Understanding the Patient Experience: A Conceptual Framework. J Patient Exp [Internet]. 2020 Dec [cited 2025 Aug 10];7(6):906. Available from: https://pmc.ncbi.nlm.nih.gov/articles/PMC7786717/

